# Maternal obesity, interpregnancy weight changes and congenital heart defects in the offspring: a nationwide cohort study

**DOI:** 10.1101/2023.02.12.23285811

**Authors:** Gitte Hedermann, Ida N Thagaard, Paula L Hedley, Lone Krebs, Christian M Hagen, Thorkild I A Sørensen, Michael Christiansen, Charlotte K Ekelund

## Abstract

**Background:** Maternal obesity has been positively associated with increased risk of congenital heart defects in the offspring. However, none of the large studies have included the considerable proportion of congenital heart defects that are identified due to improvements in prenatal diagnostics and terminated in pregnancy. The mechanism behind the association is poorly understood, and a relation to interpregnancy weight changes is to be investigated.

**Objectives:** To evaluate the association between maternal obesity and congenital heart defects in the offspring when including all pregnancies and to investigate if interpregnancy weight change between the first and second pregnancy influences risk of fetal congenital heart defects.

**Study Design:** A nationwide cohort study of all singleton pregnancies in Denmark from 2008 to 2018. All data on maternal and offspring characteristics were retrieved from the Danish Fetal Medicine Database. The database included data on postnatal diagnoses of congenital heart defects in live births and prenatal diagnoses of congenital heart defects from ultrasound examinations during pregnancy resulting in live birth, stillbirth, spontaneous abortion after gestational week 12 or termination of pregnancies after gestational week 12. As this cohort encompassed all pregnancies over a 10-year period, it was possible for women to experience multiple pregnancies. Congenital heart defects and severe congenital heart defects were grouped according to European Surveillance of Congenital Anomalies’ definitions. Children or fetuses with chromosomal aberrations were excluded. Relative risks were calculated using log-linear Poisson models for congenital heart defects overall, severe congenital heart defects and for five of the most prevalent subtypes of congenital heart defects.

**Results:** Of the 547 178 pregnancies included in the cohort, 5 498 had congenital heart defects (1.0%). Risk of congenital heart defects became gradually higher with higher maternal BMI; for BMI 30-34.9 kg/m^2^, adjusted relative risk = 1.23 (95% confidence interval 1.12-1.36), for BMI 35-39.9 kg/m^2^, adjusted relative risk = 1.26 (95% confidence interval 1.09-1.46) and for BMI ≥ 40 kg/m^2^, adjusted relative risk = 1.81 (95% confidence interval 1.50-2.15). Data was adjusted for maternal age, smoking status and year of estimated due date. The same pattern was seen for the subgroup of severe congenital heart defects. Among the atrioventricular septal defects (n = 245), a particularly strong association with maternal BMI ≥ 40 kg/m^2^ was seen, adjusted relative risk = 4.19 (95% confidence interval 2.13-7.42). 107 627 women were identified with their first and second pregnancies in the cohort. Interpregnancy BMI change was positively, albeit not statistically significant, associated with risk of congenital heart defects in the second pregnancy when adjusting for maternal age and BMI, with an adjusted relative risk = 1.27 (95% confidence interval 0.96-1.64) among persons with a BMI increase of ≥ 4 kg/m^2^.

**Conclusions:** When including both pre- and postnatally diagnosed congenital heart defects, this study showed a positive dose-response association between maternal BMI and risk of congenital heart defects in the offspring. However, only a non-significant trend was seen between interpregnancy BMI changes and risk of congenital heart defects in the second pregnancies.

**Condensation:** Tweetable statement: The risk of fetal congenital heart defect is associated with high maternal BMI, and it may also be affected by a substantial weight gain between pregnancies.

**AJOG at a Glance:** A. Why was this study conducted?

- None of the large cohort studies on the association between maternal BMI and congenital heart defects (CHDs) have included the proportion of CHDs that are identified in terminated pregnancies.
- No previous studies have investigated the association between interpregnancy weight changes and CHDs.

B. What are the key findings?

- Risk of CHDs became gradually higher with higher maternal BMI when including live births, stillbirths, abortions and terminated pregnancies.
- A trend was found between interpregnancy weight gain and risk of CHDs.

C. What does this study add to what is already known?

- The association between high maternal BMI and risk of CHDs are not caused by a lower detection rate of fetal CHDs in pregnant women with obesity.

## Introduction

Obesity among women of reproductive age has been increasing over the last three decades.^1^ Centers for Disease Control and Prevention estimated that 40% of women aged 20-39 years old in the United States were obese (body mass index [BMI] ≥ 30 kg/m^2^) in 2017-2018.^2^ Maternal obesity is a risk factor for adverse pregnancy outcomes as well as for long-term health consequences for both the mother and child.^3,4^ Furthermore, maternal obesity is positively associated with risk of having a child with congenital malformations.^5^

Congenital heart defects (CHDs) remain the leading cause of infant death from congenital malformations in the United States.^6^ Believed to be the most common congenital malformations, CHDs have a global prevalence of nine per 1000 live births with geographical differences.^7^ The causes of CHDs are unknown in most cases, but are associated with maternal chronic conditions, viral infections and fetal exposures to teratogenic drugs.^8–10^ Conflicting evidence of an association between maternal age and fetal CHDs persist, but a recent large European study found a positive association between both young- and older maternal age and total prevalence of CHDs.^11^ With improvements in genetic and genomic analytical techniques an increasing number of genetic associations/causes have been identified in up to 30% of the cases.^12^

The association between maternal obesity and infants born with congenital malformations has been reported to include CHDs. However, none of the large studies have included the proportion of CHDs that are identified and terminated in pregnancy. During the last two decades, the prenatal identification of CHDs have increased dramatically, consequently, an analysis of the association between maternal risk factors and CHDs should include data on prenatally identified cases. Meta-analyses suggest a moderate association between maternal obesity (BMI ≥ 30 kg/m^2^) and CHDs in the offspring with an odds ratio (OR) = 1.2 (95% confidence interval [CI] 1.1–1.2)^13^ or an OR = 1.3 (95% CI 1.2–1.4).^14^ A recent systematic review on the topic demonstrated great heterogeneity among the studies with respect to design, exposure definition, outcome definition, choice of covariates, and only populations of Northern European or Chinese descent were examined to a reasonable extent.^15^

Some studies have found an association between maternal interpregnancy BMI changes and adverse pregnancy- and perinatal outcomes that were linearly related to the amount of weight gain.^16,17^ So far, a few small studies suggest that this might be relevant for certain congenital malformations (spina bifida, gastroschisis and oral cleft),^16,18–20^ however, no data is available in these studies for fetal CHDs. It is of great importance to identify any modifiable risk factors for CHDs. If weight gain, defined as interpregnancy BMI change, is associated with CHDs, this could be added to the etiology of the association and the justification of preventive initiatives as to stress weight stability, and for some women weight loss.

This study aims to assess the risk of fetal CHDs, severe CHDs or five of the most frequently identified subtypes of CHDs according to early-pregnancy BMI. The study population comprises all CHDs found among live births, stillbirths, abortions and terminated pregnancies in Denmark. The study also investigates if changes in maternal BMI from the beginning of the first pregnancy to the beginning of the second pregnancy were associated with risk of CHDs in the second pregnancy.

## Materials and Methods

This cohort study was performed on prospectively collected data retrieved from a nationwide cohort based on The Danish Fetal Medicine Database.^21^ The Danish Fetal Medicine Database includes data on pregnancies with prenatal screening results from all obstetric and gynecological departments in Denmark from January 1, 2008.^21^ All women in Denmark are offered a first trimester screening for chromosomal abnormalities (gestational week 12) and a second trimester anomaly scan, for which the uptake rate is high; 95% of pregnant women participate. The database does not include data on outcomes of pregnancies before the first trimester scan. The primary source of information is the local fetal medicine databases used nationwide. The Danish Fetal Medicine Database includes data on maternal characteristics including weight and height, data from ultrasound examinations and pregnancy outcomes.^21^ Furthermore, the database includes data from other Danish registers: the Danish Cytogenetic Central Register,^22^ the Danish National Patient Register,^23^ and the Danish Medical Birth Register.^24^ All Danish residents are assigned a unique personal identification number enabling linkage of data between national registers and other data sources.^25^ In Denmark, health care is free and and it is standard practice to offer genetic testing by chorionic villus sampling or amniocentesis when a CHD is diagnosed prenatally.^26^ The prenatal detection rate of major CHD is high and the majority of parents opt for further testing.^27,28^ A gradual transition from conventional karyotyping to chromosomal microarray was observed over the course of the study period. Postnatal genetic testing primarily by chromosomal microarray is performed in all children with syndromic suspicion. The Danish Fetal Medicine database is updated once a year with information on postnatal diagnosed congenital malformations and karyotypes.^21^ The International Classification of Diseases, 10th revision code system (ICD-10) is used to code malformations in the fetus and in the infant.^21^

The cohort included singleton pregnancies in Denmark with an estimated due date, from ultrasound scan, between June 1, 2008 and June 1, 2018. Each woman can have multiple pregnancies during the study period. Pregnancies with a fetus or child with a chromosomal aberration were excluded from the cohort. Only pregnant women with a registered early-pregnancy weight and a height from 120 through 200 cm were included. Maternal BMI was calculated as weight in kilograms divided by the square of the height in meters (kg/m^2^) and BMI values are reported in that unit. Extreme observations defined as BMI < 12 and BMI > 60 were excluded to avoid registration errors.

We identified fetuses and infants with CHDs (Table S1) by using either the prenatal or/and the postnatal diagnoses. In all live births, postnatal CHD diagnoses were considered gold standard. The CHD diagnoses were defined by the European Surveillance of Congenital Anomalies (EUROCAT)^29^ (Table S1) and severe CHDs (EUROCAT, Table S1). Severe CHDs include the following 17 diagnoses: truncus arteriosus, double outlet right ventricle, transposition of the great arteries (TGA), univentricular heart (UVH), atrioventricular septum defect (AVSD), Tetralogy of Fallot (ToF), pulmonary atresia, tricuspid valve stenosis, Ebstein’s anomaly, hypoplastic right heart syndrome, aortic valve stenosis, mitral valve stenosis, mitral insufficiency, hypoplastic left heart syndrome, coarctation of the aorta (CoA), aortic atresia, and total anomalous pulmonary venous return. Irrespective of the number of CHD diagnoses in a particular patient, the patient was only registered once as having CHDs. Furthermore, offspring with five of the most frequent subtypes of severe CHDs were identified (Table S2). These were ranked as defined by Lytzen et al.^27^ with the most severe first (UVH > TGA > AVSD > CoA > ToF). If offspring had combinations of these subtypes, they were only registered once with the most severe diagnosis. ICD-10 codes for severe CHDs have been validated against hospital records with very good agreement in the Danish National Patient Register.^30^ Prenatal diagnoses of 12 severe CHDs have been shown to have a very high diagnostic precision.^28^

For the calculations of interpregnancy BMI changes, a sub-cohort including women with first and second pregnancies and more than 50 weeks between estimated due dates was compiled. Maternal height was defined as the height registered in the first pregnancy. Interpregnancy BMI changes were calculated as the difference between BMI at the beginning of the first and BMI at the beginning of the second pregnancy. Differences were categorized into six groups < −2; −2 to < −1; −1 to < 1; 1 to < 2; 2 to < 4; and ≥4 BMI units. The category −1 to < 1 was defined as stable weight and used as reference.

Associations between maternal BMI and offspring risk of CHDs were calculated as risk ratios (RRs) with 95% CI using log-linear Poisson regression models. The models were adjusted for maternal age (<20, 20–24, 25–29, 30–34, 35–39, ≥40 years), maternal smoking status (yes/no/stopped), and year of estimated due date (1-year groups). The model was not adjusted for potential multiple pregnancies for each woman during the study period. Associations between interpregnancy BMI changes and risk of CHDs were calculated as RRs with 95% CI using log-linear Poisson regression models. Maternal BMI in first pregnancy and maternal age at second pregnancy were considered possible confounders and were adjusted for in the multivariate model. Statistical analyses were run in R version 4.2.1. Statens Serum Institut has approval from the Danish Data Protection Agency to conduct register-based studies, and the project has been approved (journal no. 19/03354 and 20/09279).

## Results

The study cohort consisted of 547 178 singleton pregnancies with estimated due dates between June 1, 2008 and June 1, 2018 when pregnancies with chromosomal aberrations and missing data were excluded as detailed in Figure 1. A total of 5 498 (1.0%) offsprings had CHDs. Of these 1 227 were defined as severe CHDs (0.2%). Clinical and demographic data are available in Table 1. Information on parity was not registered and therefore missing in the dataset from 2008-2011 corresponding to 25% of the pregnancies. The study cohort comprised 534 478 live births (97.7%), 1 623 stillbirths (0.3%), 5 073 abortions or terminated pregnancies (0.9%), and 6 004 pregnancies with missing outcomes (1.1%). In total, 35% of the women had an early-pregnancy BMI ≥ 25, and 13% were obese (BMI ≥ 30). The distribution of maternal BMI among the different groups and covariates can be seen in Table 1.

**Figure 1.**
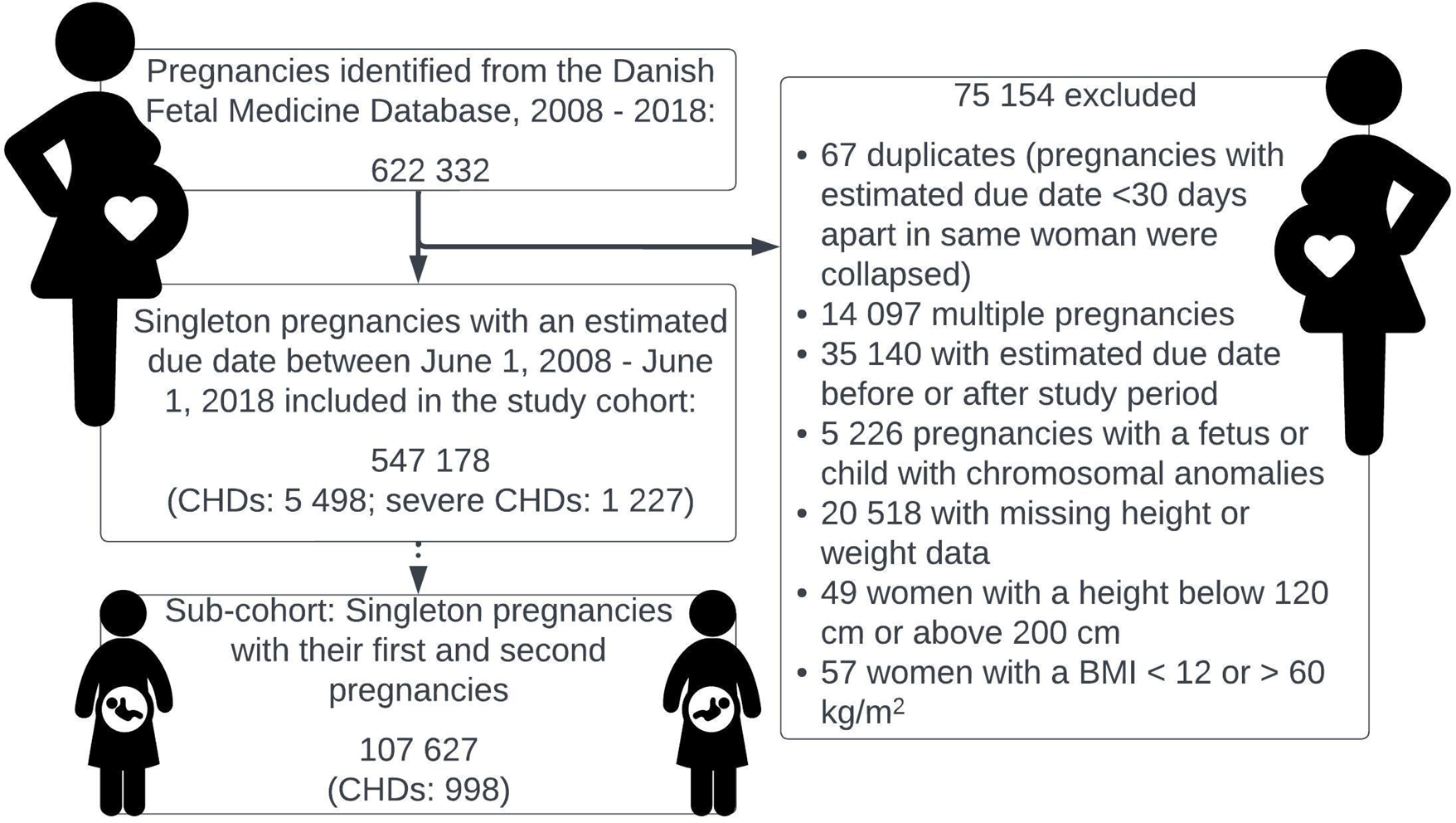
Flowchart. Abbreviations: BMI, body mass index; CHDs, congenital heart defects

**Table 1.** Maternal, pregnancy and offspring characteristics in singleton pregnancies in Denmark 2008-2018.

| Characteristics | Pregnancies with<br>congenital heart defects<br>(EUROCAT), n = 5 498 | Severe congenital<br>heart defects<br>(EUROCAT), n = 1 227 | Pregnancies without<br>congenital heart defects,<br>n = 541 680 |
| --- | --- | --- | --- |
| Body mass index, kg/m<sup>2</sup> | n (%) | n (%) | n (%) |
| < 18.5 | 371 (6.8) | 79 (6.4) | 35 792 (6.6) |
| 18.5-<25 | 2 968 (54.0) | 656 (53.5) | 318 498 (58.8) |
| 25-<30 | 1 299 (23.6) | 289 (23.6) | 117 372 (21.7) |
| 30-<35 | 531 (9.7) | 123 (10.0) | 45 927 (8.5) |
| 35-<40 | 203 (3.7) | 53 (4.3) | 16 633 (3.1) |
| ≥40 | 126 (2.3) | 27 (2.2) | 7 458 (1.4) |
| Maternal age at<br>conception, years |  |  |  |
| <20 | 125 (2.3) | 31 (2.5) | 9 371 (1.7) |
| 20-24 | 773 (14.1) | 174 (14.2) | 70 701 (13.1) |
| 25-29 | 1 871 (34.0) | 404 (32.9) | 184 442 (34.1) |
| 30-34 | 1 745 (31.7) | 403 (32.8) | 181 879 (33.6) |
| 35-39 | 843 (15.3) | 188 (15.3) | 81 370 (15.0) |
| ≥40 | 141 (2.6) | 27 (2.2) | 13 917 (2.8) |
| Ethnicity |  |  |  |
| Caucasian | 5 056 (92.0) | 1 134 (92.4) | 492 378 (90.9) |
| Non-Caucasian | 315 (5.7) | 73 (5.9) | 35 846 (6.6) |
| Missing | 127 (2.3) | 20 (1.6) | 13 456 (2.5) |
| Smoking status |  |  |  |
| No | 4 724 (85.9) | 1 080 (88.0) | 478 823 (88.4) |
| Yes | 627 (11.4) | 115 (9.4) | 50 238 (9.3) |
| Stopped | 92 (1.7) | 20 (1.6) | 8 765 (1.6) |
| Missing | 55 (1.0) | 12 (1.0) | 3 854 (0.7) |
| Conception |  |  |  |
| Spontaneous | 4 950 (90.0) | 1 117 (91.0) | 497 936 (91.9) |
| Assisted | 463 (8.4) | 97 (7.9) | 36 516 (6.8) |
| Unknown | 85 (1.5) | 13 (1.1) | 7 228 (1.3) |
| Parity |  |  |  |
| 0 | 2 046 (37.2) | 416 (33.9) | 181 065 (33.4) |
| 1 | 1 571 (28.6) | 367 (29.9) | 153 479 (28.3) |
| 2 | 540 (9.8) | 133 (10.8) | 53 959 (10.0) |
| ≥3 | 203 (3.7) | 38 (3.1) | 16 653 (3.1) |
| Missing | 1 138 (20.7) | 273 (22.2) | 136 524 (25.2) |
| Year of estimated due date |  |  |  |
| 2008 | 216 (3.9) | 57 (4.6) | 25 266 (4.7) |
| 2009 | 477 (8.7) | 126 (10.3) | 56 468 (10.4) |
| 2010 | 550 (10.0) | 131 (10.7) | 57 687 (10.7) |
| 2011 | 528 (9.7) | 120 (9.8) | 53 630 (9.9) |
| 2012 | 534 (9.7) | 137 (11.2) | 53 512 (9.9) |
| 2013 | 508 (9.2) | 120 (9.8) | 51 331 (9.5) |
| 2014 | 590 (10.7) | 141 (11.5) | 52 609 (9.7) |
| 2015 | 642 (11.7) | 137 (11.2) | 54 215 (10.0) |
| 2016 | 607 (11.0) | 110 (9.0) | 57 402 (10.6) |
| 2017 | 543 (9.9) | 101 (8.2) | 56 569 (10.4) |
| 2018 | 303 (5.5) | 47 (3.8) | 22 991 (4.2) |
| Outcome pregnancy |  |  |  |
| Live births | 5 187 (94.3) | 1 011 (82.4) | 529 291 (97.7) |
| Stillbirths | 41 (0.7) | 26 (2.1) | 1 582 (0.3) |
| Abortions and terminated pregnancies after gestational week 12 | 245 (4.5) | 175 (14.3) | 4 828 (0.9) |
| Missing | 25 (0.5) | 15 (1.2) | 5 979 (1.1) |
| Offspring sex |  |  |  |
| Male | 2 771 (50.4) | 624 (50.9) | 271 571 (50.1) |
| Female | 2 416 (43.9) | 387 (31.5) | 257 720 (47.6) |
| Unknown | 311 (5.7) | 216 (17.6) | 12 389 (2.3) |

Abbreviations: EUROCAT, European Surveillance of Congenital Anomalies (diagnoses listed in Table

Maternal overweight was associated significantly with higher risk of CHDs in the offspring increasing with higher maternal BMI: for BMI 25-29.9, adjusted relative risk (aRR) = 1.17 (95% CI 1.09-1.25), for BMI 30-34.9, aRR = 1.23 (95% CI 1.12-1.36), for BMI 35-39.9, aRR = 1.26 (95% CI 1.09-1.46) and for BMI ≥ 40, aRR = 1.81 (95% CI 1.50-2.15) compared to women with an early-pregnancy BMI 18.5-24.9 (Table 2). Similar results were seen for maternal BMI and severe CHDs: for BMI 25-29.9, aRR = 1.20 (95% CI 1.04-1.38), for BMI 30-34.9, aRR = 1.28 (95% CI 1.04-1.56), for BMI 35-39.9, aRR = 1.47 (95% CI 1.09-1.96) and for BMI ≥ 40, aRR = 1.85 (95% CI 1.23-2.67) compared to women with an early-pregnancy BMI 18.5-24.9 (Table 2). A slightly higher risk of CHDs, when only including live births compared to all pregnancies, was seen for maternal BMI ≥ 30 (aRR 1.24-1.87; data available in Table S3).

**Table 2.** Relative risks of congenital heart defects by maternal BMI in 547 178 offsprings (live births, stillbirths, abortions and terminated pregnancies), Denmark 2008-2018.

| Maternal<br>BMI (kg/m<sup>2</sup>) | No CHDs | CHDs |  |  |  |  | Severe CHDs |  |  |  |  |
| --- | --- | --- | --- | --- | --- | --- | --- | --- | --- | --- | --- |
|  | n = 541 680 | n = 5 498 |  |  |  |  | n = 1 227 |  |  |  |  |
|  | Total | Total | Crude<br>RR | 95% CI | aRR* | 95% CI | Total | Crude RR | 95% CI | aRR* | 95% CI |
| < 18.5 | 35 800 | 371 | 1.11 | 1.00-1.24 | 1.07 | 0.95-1.19 | 79 | 1.07 | 0.84-1.34 | 1.02 | 0.79-1.30 |
| 18.5 - 24.9 | 318 489 | 2 968 | 1.00 | ref | 1.00 | ref | 656 | 1.00 | ref | 1.00 | ref |
| 25 - 29.9 | 117 373 | 1 299 | 1.19 | 1.11-1.27 | 1.17 | 1.09-1.25 | 289 | 1.19 | 1.04-1.37 | 1.20 | 1.04-1.38 |
| 30 - 34.9 | 45 926 | 531 | 1.24 | 1.13-1.36 | 1.23 | 1.12-1.36 | 123 | 1.30 | 1.07-1.57 | 1.28 | 1.04-1.56 |
| 35 - 39.9 | 16 635 | 203 | 1.31 | 1.13-1.50 | 1.26 | 1.09-1.46 | 53 | 1.55 | 1.15-2.02 | 1.47 | 1.09-1.96 |
| ≥ 40 | 7 457 | 126 | 1.80 | 1.50-2.14 | 1.81 | 1.50-2.15 | 27 | 1.76 | 1.17-2.52 | 1.85 | 1.23-2.67 |

Abbreviations: aRR, adjusted relative risk; BMI, body mass index; CHD, congenital heart defects; CI, confidence interval; RR, relative risk.
\*aRR adjusted for maternal age at conception, smoking status (yes/no/stopped) and year of estimated due date comparing CHD risk in women with early-pregnancy BMI < 18.5 kg/m<sup>2</sup> or BMI ≥ 25 kg/m<sup>2</sup> with women with early-pregnancy BMI in the normal range (BMI 18.5-24.9 kg/m<sup>2</sup>).

The proportion of CHD cases with one of the five specific CHD diagnoses were as follows: univentricular heart (UVH; 4.4%), transposition of the great arteries (TGA; 3.2%), atrioventricular septum defect (AVSD; 4.5%), coarctation of the aorta (CoA; 4.6%), and Tetralogy of Fallot (ToF; 2.3%). The association between maternal BMI and these specific five CHD diagnoses are shown in Figure 2. No significant associations were seen for UVH, TGA, CoA and ToF. However, maternal BMI ≥ 30 was associated with a significantly higher risk of AVSD in the offspring (Figure 2).

**Figure 2.**
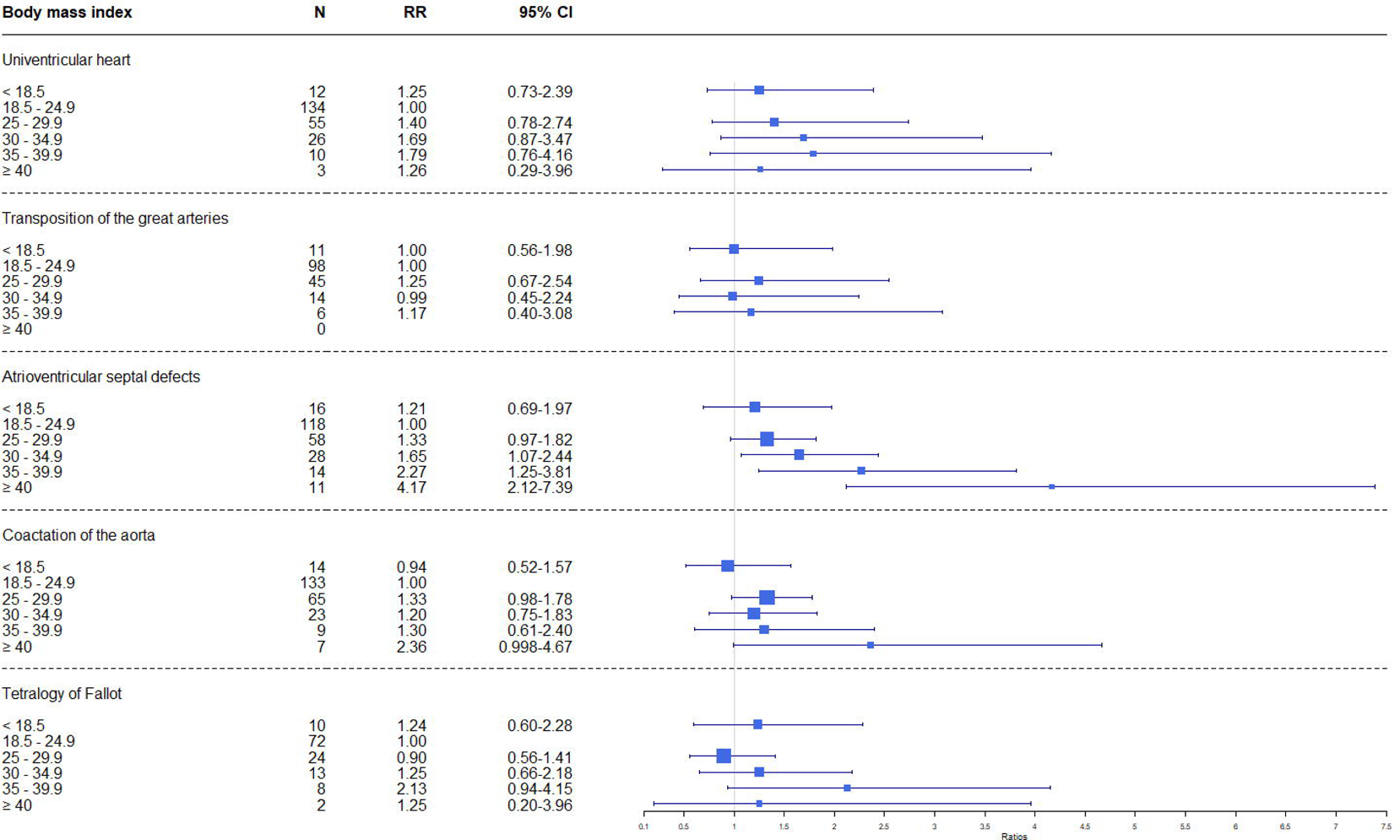
Crude relative risks for five subtypes of congenital heart defects by maternal BMI, singleton pregnancies in Denmark 2008-2018. Body mass index 18.5-24.9 kg/m^2^ was considered as the normal range and used as a reference. Abbreviations: CHDs, congenital heart defects; CI, confidence interval; RR, relative risk; N, number of events

The sub-cohort included 107 627 women who had a first and second consecutive singleton pregnancies between June 1, 2008 and June 1, 2018 (Table 3). Mean BMI in first pregnancy was 24.9 vs. 25.8 in second pregnancy. In total, 998 had an offspring with CHDs (0.9%) in their second pregnancy. The prevalence of CHDs in the second pregnancy became higher with increased weight gain between pregnancies (0.8% to 1.3%). Increase in maternal BMI between pregnancies were significantly associated with higher risk of CHDs in second pregnancy. However, when adjusted for maternal age in the second pregnancy and maternal BMI in the first pregnancy, only a trend remained towards higher risks of any CHDs with weight gain between pregnancies (Table 3).

**Table 3.** Relative risks of congenital heart defects in second pregnancy by interpregnancy maternal BMI changes in 107 627 women with two consecutive singleton pregnancies, Denmark 2008-2018.

| BMI change from 1st to 2nd pregnancy |  | 1st pregnancy | 2nd pregnancy | Risk of CHDs in 2nd pregnancy |  |  |  |  |  |
| --- | --- | --- | --- | --- | --- | --- | --- | --- | --- |
| Units, kg/m<sup>2</sup> | No. of women with 1st and 2nd pregnancy (%) | Mean BMI | Mean BMI | n/N | Prevalence of CHDs (%) | Crude RR | 95% CI | aRR* | 95% CI |
| <-2 | 5 941 (5.5) | 27.3 | 24.3 | 50/5 891 | 0.8 | 0.95 | 0.70-1.26 | 0.84 | 0.61-1.12 |
| -2 to <-1 | 11 154 (10.4) | 24.2 | 22.8 | 90/11 064 | 0.8 | 0.91 | 0.72-1.14 | 0.88 | 0.70-1.09 |
| -1 to <1 | 54 154 (50.3) | 22.8 | 22.9 | 479/53 675 | 0.9 | 1.00 | ref | 1.00 | ref |
| 1 to <2 | 16 940 (15.7) | 23.8 | 25.2 | 156/16 784 | 0.9 | 1.04 | 0.87-1.24 | 1.01 | 0.84-1.21 |
| 2 to <4 | 14 010 (13.0) | 25.0 | 27.8 | 155/13 855 | 1.1 | 1.25 | 1.04-1.50 | 1.17 | 0.97-1.40 |
| ≥ 4 | 5 428 (5.0) | 26.3 | 31.4 | 68/5 360 | 1.3 | 1.42 | 1.09-1.81 | 1.27 | 0.96-1.64 |
| Total | 107 627 | 24.9 | 25.8 | 998/106 629 | 1.0 |  |  |  |  |

Abbreviations: aRR, adjusted relative risk; BMI, body mass index; CHDs, congenital heart defects; CI, confidence interval; n, number of women with their first pregnancy without CHDs and second pregnancy with CHDs in the offspring; N, number of women with first and second pregnancies without CHDs; RR, relative risk.
\*aRR adjusted for maternal age at conception at second pregnancy and early-pregnancy BMI at 1st pregnancy.

## Comment

### Principal Findings

When including both pre- and postnatally diagnosed CHDs, this study showed a dose-response association between high maternal BMI and risk of CHDs in the offspring. However, only a non-significant trend was seen between interpregnancy BMI changes between first and second pregnancies and risk of fetal CHDs in the second pregnancies.

### Results in the Context of What is Known

This prospective nationwide cohort included 547 178 live births, stillbirths, abortions and terminated pregnancies in Denmark from 2008 to 2018 and showed that maternal overweight and obesity were significantly associated with a moderately higher risk of CHDs (aRR = 1.17-1.81) and severe CHDs (aRR = 1.20-1.85) in the offspring compared to women with a BMI in the normal range (BMI 18.5-24.9). The study validates previous findings of a positive association between high maternal BMI and risk of CHDs in the offspring^13,14,31,32^ and thereby rejects any hypothesis of this association to be caused by lower prenatal detection rates of CHDs in pregnant women with obesity. Most other studies have been limited to live births.^15,31^ A large nationwide Swedish study with two million live born children found a positive association between high maternal BMI and CHDs (BMI 30-34.9: OR = 1.2, 95% CI 1.2–1.3; BMI ≥ 40: OR = 1.6, 95% CI 1.4–1.8).^31^ When restricting our analysis to live births (Table S3), we found significant associations of a similar magnitude between maternal BMI ≥ 25 and CHDs. Results from five specific subtypes of CHDs (UVH, TGA, AVSD, CoA and ToF) showed no significant association with maternal BMI except for AVSD (Figure 2), which was significantly associated with increased risks when maternal BMI ≥ 30 (BMI 30 to ≥ 40: aRR = 1.65-4.19). Persson et al. found a non-significant trend between maternal BMI and AVSD in women with BMI ≥ 30,^5^ and the same pattern was observed in other studies.^9,33,34^ The lack of statistical significance in other studies might be due to few cases in each BMI group or different study designs. We examined the interpregnancy weight changes between the first and second pregnancies in 107 627 women and found a trend towards higher risks of CHDs in second pregnancies preceded by a substantial BMI gain but this was not significant when data was adjusted for maternal BMI in first pregnancy and maternal age in second pregnancy (Table 3) as a result of the correlation between maternal BMI and interpregnancy weight changes. The investigations of the association between interpregnancy weight changes and risk of fetal CHDs are sparse.^16^ A few studies have looked at other congenital malformations and found a RR = 2.3 for isolated cleft palate when maternal BMI increased ≥ 3 units,^20^ a positive association between spina bifida and interpregnancy BMI gain,^18^ and a significant decrease in OR = 0.62 (95% CI 0.42-0.94) for gastroschisis when maternal BMI increased with ≥ 3 units.^19^

### Clinical Implications

Lifestyle interventions to reduce risk of fetal CHDs have been suggested to be introduced before or between pregnancies.^35^ Although our results only indicate a trend between interpregnancy weight changes and risk of CHDs, it does not dismiss a positive effect of weight reduction in women with obesity before pregnancy. In a setting with pre-pregnancy counseling, it is still important to advise women about the importance of BMI as a risk factor for congenital malformations,^5^ obstetric and perinatal complications.^3,4^

### Reseach Implications

Prenatal detection rates of congenital malformations decreases with increasing maternal BMI since image quality is lower in women with obesity.^36,37^ Consequently, studies only including live births could be biased towards a higher postnatal prevalence of CHDs in women with obesity if severe fetal CHDs were not diagnosed and possibly terminated during pregnancy (Table 1). Thus, this phenomenon may at least partly explain a positive association between high BMI and prevalence of CHDs in live borns.^5^ Prenatal detection rates of CHDs have improved substantially in the last decades, and in countries with prenatal screening with high detection rate of CHDs as in Denmark,^28^ it is more important to include terminated pregnancies in prevalence and association studies as the rate of terminated pregnanciess with the most severe CHDs likely will be higher as seen in Table 1.

This study confirms the association between high maternal BMI and risk of fetal CHDs. Knowledge about the etiology is still limited^35^ and future research should focus on combinations of other related maternal metabolic disorders linked to insulin resistance as suggested in a recent review.^15^

### Strengths and Limitations

The strength of this study is the large size of the cohort and prospectively, nationwide data collection including prenatal information. Some limitation must be considered. The database did not include data on pregestational diabetes, which is known to be strongly associated with CHDs, and therefore this confounder was not included as a covariate in the analyses.^15,38^ Persson et al. excluded all women with pregestational diabetes and found a moderate association similar to the results of the present study.^31^ Nor did the data include information about family history of CHDs,^39^ maternal infections or teratogenic medicine intake in pregnancy that have been associated with higher risk of CHDs.^8^ Since the study draws conclusions from register data, there is a risk of reporting bias and all CHD diagnoses are not validated against hospital records.

## Conclusions

High maternal BMI is associated with risk of fetal CHDs, and while substantial weight gain between pregnancies may impact the risk, it does not appear to have a significant effect on it.

## Data Availability

All data produced in the present study are available upon reasonable request to the authors

## Disclosures

The authors report no conflict of interest

## Funding

This study was supported by The Danish Children Heart Foundation (18-R109-A5193-26043), The A.P. Moller Foundation (19-L-0096), and Aase and Ejnar Danielsen’s Foundation (19-10-0493). This research has been conducted using the Danish National Biobank resource supported by the Novo Nordisk Foundation. The funders had no role in study design, data collection and analysis, decision to publish, or preparation of the manuscript.

## Acknowledgements

The study group wishes to thank sonographers and medical doctors, who have and continuously are collecting data for the Danish Fetal Medicine Database.

An earlier version of the article is available at a preprint server, MedRxiv (https://medrxiv.org/cgi/content/short/2023.02.12.23285811v1).

## Glossary

AVSD: atrioventricular septum defect
CHDs: congenital heart defects
CoA: coarctation of the aorta
EUROCAT: European Surveillance of Congenital Anomalies
TGA: transposition of the great arteries
ToF: Tetralogy of Fallot
UVH: univentricular heart

## Notes

### Competing Interest Statement

The authors have declared no competing interest.

### Funding Statement

This study was funded by The Danish Children Heart Foundation (18-R109-A5193-26043), The A.P. Moller Foundation (19-L-0096), and Aase and Ejnar Danielsen's Foundation (19-10-0493).

### Author Declarations

Statens Serum Institut has approval from the Danish Data Protection Agency to conduct register-based studies, and the project has been approved (journal no. 19/03354 and 20/09279).

### Summary of Updates

Updated version

