## Supplementary materials for "Maternal obesity, interpregnancy weight changes and congenital heart defects in the offspring: a nationwide cohort study"

Table S1. Definition of congenital heart defects by ICD-codes

|  | ICD-10 | Comments | Excluded diagnoses |
| --- | --- | --- | --- |
| CHDs (EUROCAT) | Q20-26 | Exclude PDA with GA<37 weeks; Exclude peripheral pulmonary artery stenosis with GA<37 weeks | Q211C (Q2111: Patent or persistent foramen ovale), Q250 if GA<37 weeks (PDA), Q254E (Q2541: Persistent right aortic arch), Q256 if GA<37 weeks (Peripheral pulmonary artery stenosis), Q261 (Persistent left superior vena cava) |
| Severe CHDs (EUROCAT) | Q200 | Truncus arteriosus |  |
|  | Q201 | Double outlet right ventricle |  |
|  | Q203 | Transposition of the great arteries (TGA) |  |
|  | Q204 | Univentricular heart (UVH) |  |
|  | Q212 | Atriventricular septal defects (AVSD) |  |
|  | Q213 | Tetralogy of Fallot (ToF) |  |
|  | Q220 | Pulmonary atresia |  |
|  | Q224 | Tricuspid valve stenosis |  |
|  | Q225 | Ebstein’s anomaly |  |
|  | Q226 | Hypoplastic right heart syndrome |  |
|  | Q230 | Aortic valve stenosis |  |
|  | Q232 | Mitral valve stenosis |  |
|  | Q233 | Mitral insufficiency |  |
|  | Q234 | Hypoplastic left heart syndrome |  |
|  | Q251 | Coarctatio aortae (CoA) |  |
|  | Q252 | Interrupted aortic arch |  |
|  | Q262 | Total anomalous pulmonary venous return |  |

Table S2. Definitions of five of the most frequently identified, perinatally as well as neonatally, subtypes of severe congenital heart defects

| ICD-10 | Diagnoses |
| --- | --- |
| DQ204  DQ234  DQ226 | Univentricular heart (UVH)  Prenatally diagnosed - hypoplastic left heart syndrome  Prenatally diagnosed - hypoplastic right heart syndrome |
| DQ203 | Transposition of the great arteries (TGA) |
| DQ212 | Atriventricular septal defects (AVSD) |
| DQ251 | Coarctatio aortae (CoA) |
| DQ213 | Tetralogy of Fallot (ToF) |

Table S3. Live births and risk of congenital heart defects

|  | No CHDs | CHDs | | | | |
| --- | --- | --- | --- | --- | --- | --- |
|  | n = 529 185 | n = 5179 | | | | |
| Maternal BMI (kg/m2) | Total | Total | Crude RR | 95% CI | Crude RR | 95% CI |
| < 18.5 | 34 835 | 351 | 1.12 | 1.01-1.25 | 1.08 | 0.96-1.21 |
| 18.5 - 24.9 | 311 250 | 2 785 | 1.00 | ref | 1.00 | ref |
| 25 - 29.9 | 114 717 | 1 224 | 1.19 | 1.11-1.27 | 1.17 | 1.09-1.25 |
| 30 - 34.9 | 44 864 | 499 | 1.24 | 1.13-1.36 | 1.24 | 1.12-1.36 |
| 35 - 39.9 | 16 230 | 197 | 1.35 | 1.17-1.56 | 1.31 | 1.12-1.51 |
| ≥ 40 | 7 289 | 123 | 1.87 | 1.55-2.23 | 1.87 | 1.55-2.24 |

Congenital heart defects defined by EUROCAT

Abbreviations: aRR, adjusted relative risk; BMI, body mass index; CHDs, congenital heart defects; CI, confidence interval; RR, relative risk.

*aRR adjusted for maternal age at conception, smoking status (yes/no/stopped) and year of estimated due date comparing CHD risk in women with early-pregnancy BMI < 18.5 kg/m^2^ or BMI ≥ 30 kg/m^2^ with women with early-pregnancy normal BMI (18.5-24.9 kg/m^2^).
